# Gene-based map of cardiovascular diseases

**DOI:** 10.64898/2026.11.11.26367883

**Authors:** Barış Kayaalp, Kerem Çil, Tayfun Özçelik

## Abstract

**Background:** Rare coding variants contribute to cardiovascular disease, but their effects across disease groups remain incompletely characterized. In particular, the influence of functionally and clinically informed variant classification and sex-specific genetic effects on gene–phenotype associations remains understudied.

**Methods:** We analyzed whole-genome sequencing data from up to 483,311 UK Biobank participants across six genetic ancestry groups and eight cardiovascular phenotype groups. Gene-burden testing integrated state-of-the-art missense classification and predicted pathogenic germline variants. We performed combined-sex and sex-stratified analyses, followed by cross-ancestry meta-analysis. We additionally adjusted for cardiometabolic traits in sensitivity analyses, estimated penetrance among qualifying-variant carriers, and assessed genetic prevalence across eight ancestry groups.

**Results:** We identified 86 exome-wide significant gene–phenotype associations: 40 established and 46 novel. Novel associations included *EHMT2* with atrial fibrillation, *P2RX1* with arrhythmias, *P2RY4* with complete atrioventricular block, and *NOSTRIN* with atherosclerotic heart disease in females. After cardiometabolic adjustment, 60 associations retained exome-wide significance and 18 additional associations emerged, including *CDH11*, *PARVA*, and *ADAMDEC1* with atherosclerosis-related phenotypes.

**Conclusions:** These findings identify novel cardiovascular gene–phenotype associations implicating purinergic signaling, endothelial nitric oxide regulation, and vascular wall biology. Variant selection shapes penetrance estimates and the interpretation of inherited susceptibility.

## Introduction

Inherited variation contributes to cardiovascular disorders through effects on cardiac electrical activity, myocardial and valvular function, vascular structure, tissue perfusion, and haemostasis. Genetic effects on lipid metabolism further contribute to susceptibility to atherosclerotic cardiovascular disease. Human genetics has provided foundational causal insights across these domains. Mutations in *KCNH2* and *SCN5A* established ion-channel dysfunction as a cause of inherited arrhythmia^1,2^, while *MYH7* mutations associated with the familial hypertrophic cardiomyopathy^3^. *FBN1* mutations linked extracellular-matrix defects to inherited aortopathy^4^, *NOTCH1* mutations established a genetic basis for aortic-valve disease^5^, and the *F5* Leiden variant connected activated protein C resistance to inherited thrombotic risk^6^. *NOTCH3* mutations identified a cause of inherited cerebral small-vessel disease^7^, while *PCSKS* variants linked lower LDL cholesterol to reduced coronary heart disease risk^8^. Despite these advances, the gene-level contribution of rare coding variation across the broader spectrum of cardiovascular disease remains incompletely characterized.

Population-scale sequencing enables systematic investigation of rare coding variation, although low allele counts often limit the power of single-variant association tests^9,10^. Gene-based burden testing aggregates qualifying variants within each gene, enabling detection of associations driven by multiple rare alleles. Its power depends critically on variant selection: inclusion of neutral alleles dilutes signals, whereas exclusion of functionally consequential alleles reduces sensitivity^11^. Missense variants pose a particular challenge because their functional effects and clinical significance are often uncertain^12^. Combining clinically informed classification with predictions grounded in functional evidence may therefore improve the selection of variants for gene-based discovery.

Sex and ancestry add further complexity. Genetic effects may differ between females and males, while allele frequencies and rare-variant spectra vary across ancestries^9,13^. Combined and stratified analyses can therefore provide complementary views of gene– phenotype associations. For coronary, cerebrovascular and other atherosclerotic outcomes, cardiometabolic risk factors also need to be considered when interpreting associations. In parallel, assessing the population distribution of qualifying genotypes and disease expression among carriers can help place gene-based discoveries in a clinically relevant context.

Here, we performed clinically and functionally informed rare-variant gene-burden analyses using whole-genome sequencing and linked clinical data from up to 483,311 UK Biobank participants across eight cardiovascular phenotype groups: aneurysm and dissection; arrhythmia and conduction disease; cardiomyopathy and heart failure; valvular disorders; embolism and thrombosis; cerebrovascular disease; coronary and ischaemic heart disease, including myocardial infarction; and atherosclerotic disease. We conducted combined-sex and sex-stratified analyses across six genetic ancestry groups, followed by cross-ancestry meta-analysis. We used AAVC for ACMG/AMP-informed variant classification^14^ and FuncVEP-CTE for functional-evidence-informed missense prioritization^15^, evaluating high-impact, ACMG-classified and predicted damaging missense masks across multiple allele-frequency thresholds. We assessed associations after additional adjustment for cardiometabolic risk factors and estimated phenotype penetrance among qualifying-variant carriers in UK Biobank. We also estimated the prevalence of qualifying genotypes across eight genetic ancestry groups, extending our framework for quantifying inherited disease predisposition^16^. Together, these analyses aimed to broaden the gene-based map of cardiovascular disease and characterize the population distribution and phenotypic expression of associated rare variation.

## Methods

### Study cohort

UK Biobank is composed of nearly half a million participants with their ages ranging from 40 to 69, with genome sequencing and linked electronic health data. Whole-genome sequencing data was available for 490,640 UK Biobank participants and its processing was described previously^17,18^, with 483,311 participants retained after quality control (Supplementary Information: Quality control).

### Variant classification

We utilized two tools for variant classification: AAVC^14^ for ACMG-based variant classification and prioritization of high-impact variants; and FuncVEP-CTE^15^ for missense predictions. We defined variant sets under four broad mask categories: ACMG-classified, damaging missense, high-impact, and synonymous variants used for inflation assessment (Supplementary Information: Variant classification).

### Phenotype definitions

For disease phenotypes, we mapped ICD-10 diagnosis codes from the UK Biobank to broader disease categories comprising aortic aneurysm or dissection, arrhythmia, cardiomyopathies and heart failure, valvular disorders, embolism and thrombosis, cerebrovascular diseases, and myocardial infarction (Supplementary Table 1). In addition to individual phenotype definitions, we constructed combined phenotype groups by aggregating related diagnoses within each major disease category. We additionally defined an atherosclerotic disease group comprising atherosclerotic phenotypes across cardiovascular vascular territories, including aortic and coronary atherosclerosis, as well as ischemic stroke.

### Statistics

We performed gene-based association analyses using REGENIE v4.1^19^ and remeta v0.11.1^20^. In the combined-sex analyses, models were adjusted for age, age squared, sex, Townsend deprivation index, and the first 20 genetic principal components (Supplementary Information: Statistics).

### Gene-prioritization

To prioritize associations, we calculated genomic inflation factor for all gene-phenotype associations using synonymous variants for each sex and allele frequency mask. We kept associations with inflation factor lower than 1.15 (Supplementary Table 2). To further prioritize the associations, we included the ones observed in more than or equal to 10 individuals.

### Calculation of penetrance, carrier frequency and genetic prevalence

Penetrance was estimated as the posterior mean under a binomial model, with 95% confidence intervals. Carrier frequencies in UK Biobank were calculated as the proportion of carriers among all analysed participants, with 95% confidence intervals. Unweighted linear regression assessed the relationship between estimated penetrance and log₁₀-transformed inverse carrier frequency.

Carrier frequency and genetic prevalence were estimated for genes associated with cardiometabolic risk or protection using allele-frequency data from the eight genetic ancestry groups represented in gnomAD: African, Admixed American, Ashkenazi Jewish, East Asian, Finnish, Middle Eastern, non-Finnish European, and South Asian. (Supplementary Information: Genetic prevalence).

### Data availability

Gene-phenotype association results produced in this study are available at https://zenodo.org/records/23687128. All other data generated or analysed during this study are included in the Supplementary Tables.

We used publicly available software’s: AAVC and FuncVEP are available at https://github.com/OzcelikLab/AAVC and https://github.com/OzcelikLab/FuncVEP. REGENIE and remeta are available at https://rgcgithub.github.io/regenie/ and https://rgcgithub.github.io/remeta/.

## Results

### Summary of gene-based association on cardiovascular traits on UK Biobank

We conducted sex-stratified gene-based burden analyses using whole-genome sequencing data from up to 483,311 UK Biobank participants across six genetic ancestry groups: African, Admixed American, Central and South Asian, East Asian, European, and other ancestries. Analyses were performed using REGENIE and followed by cross-ancestry meta-analysis using remeta. We defined multiple variant masks comprising AAVC-classified pathogenic (P), likely pathogenic (LP), and high-confidence variants of uncertain significance (VUS-H); high-impact variants; predicted damaging missense variants; and synonymous variants as a negative control. Each variant mask was evaluated across a range of allele-frequency thresholds, from a minor allele frequency of 1% to singleton variants. The analyses encompassed eight cardiovascular phenotype groups (Figure 1). To distinguish replicated from novel findings, we curated previously established gene–phenotype associations from PanelApp, OMIM, and the published literature. The resulting reference set comprised 672 genes (Supplementary Table 3). Overall, we identified 86 exome-wide significant gene–phenotype associations, of which 40 replicated previously established relationships and 46 were novel (Table 1, Supplementary Table 4).

**Figure 1:**
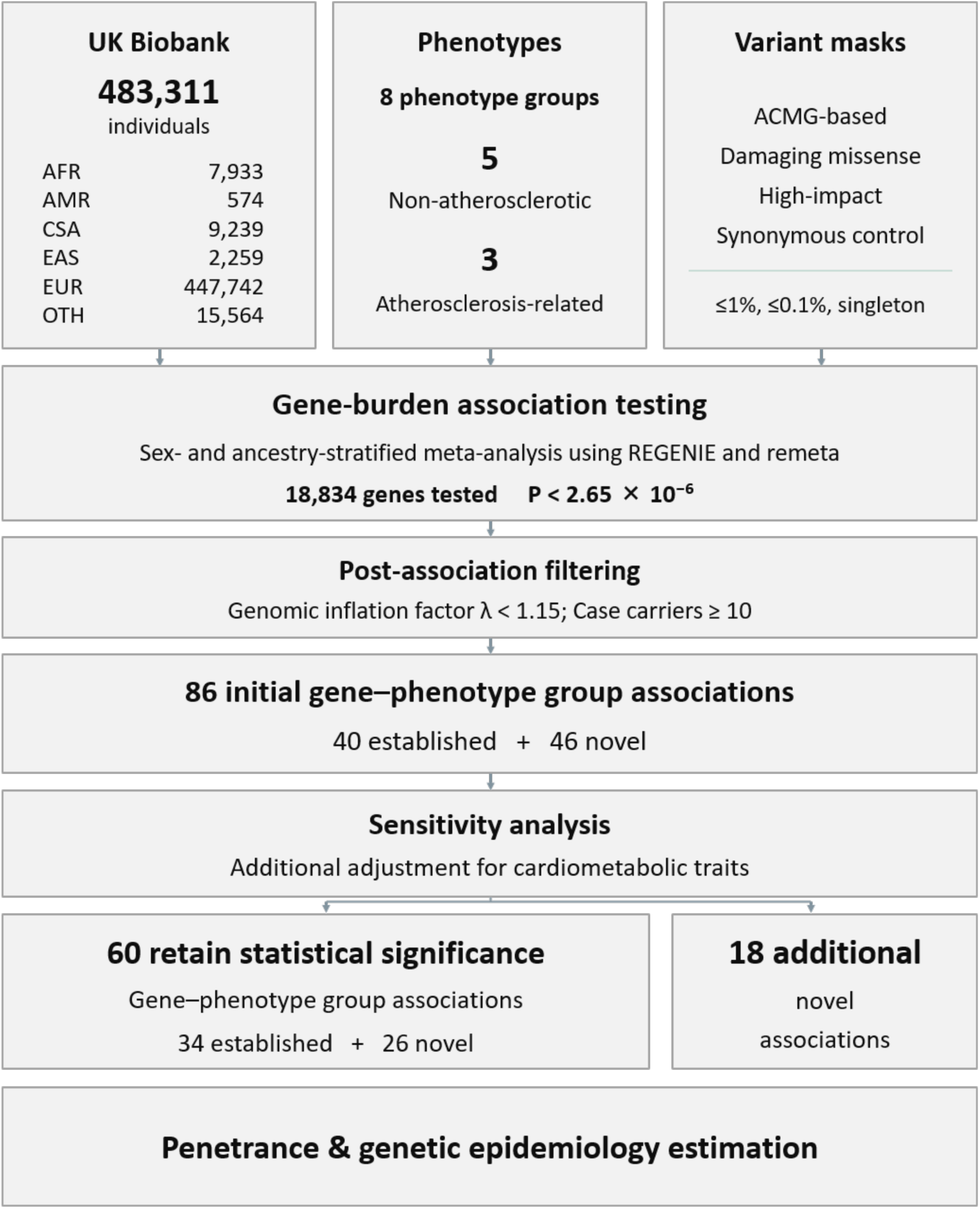
Study design. Whole-genome sequencing data from up to 483,311 UK Biobank participants were analysed across six genetic ancestry groups using gene-based burden testing with REGENIE, followed by cross-ancestry meta-analysis using remeta. Variants were prioritized using AAVC for ACMG/AMP-informed pathogenicity classification and FuncVEP-CTE for functional prediction of missense variants. Multiple variant masks and allele-frequency thresholds were evaluated across cardiovascular phenotype groups in combined-sex and sex-stratified analyses. Significant gene–phenotype associations were classified as previously established or novel using curated genetic evidence from PanelApp, OMIM, and published literature. Additional analyses included adjustment for conventional cardiometabolic risk factors, estimation of phenotype penetrance among qualifying-variant carriers, and assessment of genetic prevalence across eight gnomAD ancestry groups.

**Table 1:** Summary statistics for known, replication and novel genes.

| **Cardiovascular diseases** | **Known** | **Replication** | **Novel** |
|---|---|---|---|
| **Aortic aneurysm and dissection** | 52 | 2 | 1 |
| **Arrhythmia** | 188 | 13 | 15 |
| **Cardiomyopathy and heart failure** | 342 | 8 | 4 |
| **Embolism and thrombosis** | 30 | 5 | 4 |
| **Valve disorder** | 197 | 2 | 6 |
| **Cerebrovascular disease** | 79 | 3 | 6 |
| **Myocardial infarction** | 54 | 6 | 7 |
| **Atherosclerosis** | - | 1 | 3 |
| **Sum** | 942<sup>*</sup> | 40 | 46 |

<sup>\*</sup>: When the unique count of genes are considered the number reduces to 672.

### Gene-burden results for aortic aneurysms and dissections

In our analysis, three genes—*FBN1*, *SMAD3*, and *GOSR2*—reached exome-wide significance, all for the combined phenotype comprising all aortic aneurysms. *FBN1* and *SMAD3* are established aneurysm genes, causing Marfan syndrome and Loeys– Dietz syndrome, respectively (Supplementary Table 5, Figure 2A).

**Figure 2:**
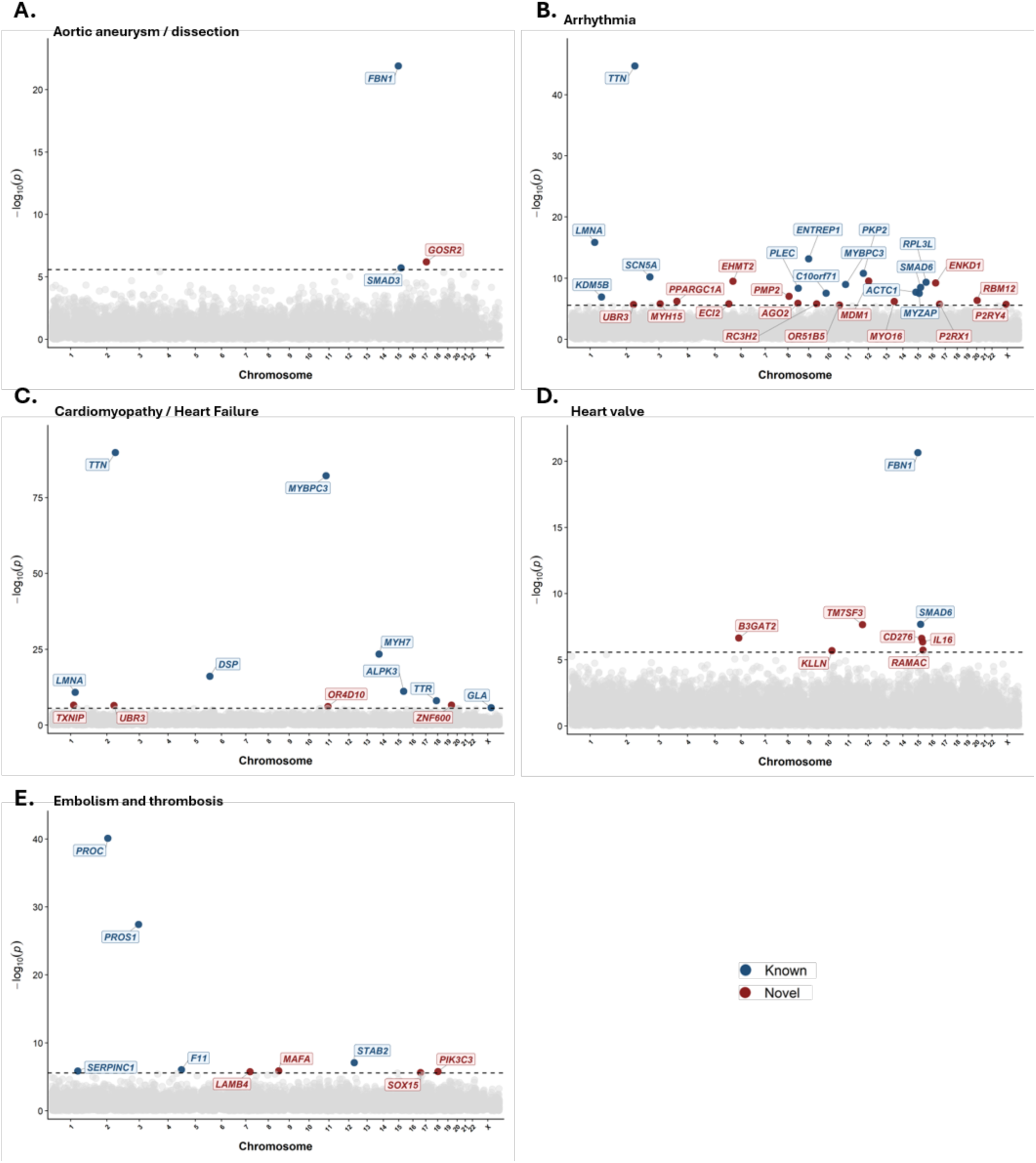
Manhattan plots for gene-based association results for non-atherosclerotic cardiovascular phenotype groups. Each point represents a gene, positioned according to its genomic location across chromosomes 1–22 and X. The vertical axis indicates the negative base-10 logarithm of the association P-value (−log₁₀P). For each gene within a phenotype group, the lowest P-value across eligible association tests was retained. Associations were restricted to analyses with at least 10 qualifying-variant carriers among cases. The horizontal dashed line indicates the Bonferroni-corrected significance threshold of P = 0.05/18,834. Previously established gene–phenotype associations are highlighted in dark blue, whereas novel associations are highlighted in dark red.

*GOSR2* encodes Golgi SNAP receptor complex member 2, a Qb-SNARE protein involved in vesicular transport between the endoplasmic reticulum and Golgi apparatus. Biallelic pathogenic variants in *GOSR2* cause recessive disorders, including progressive myoclonus epilepsy/ataxia and congenital muscular dystrophy (OMIM: 614018 and 620166). In our analysis, heterozygous carriers of P+LP *GOSR2* variants were associated with the combined aortic aneurysm phenotype in males (OR = 5.10, P = 6.47 × 10⁻⁷, λ = 0.95), with qualifying variants observed in 15 cases and 177 controls. Independent common-variant studies suggested that variants at or near *GOSR2* have been associated with aortic root diameter and left ventricular outflow tract dimensions^21,22^, as well as measures of aortic valve function^23^. In addition, a genome-wide association study of congenital heart disease identified variants within the *GOSR2* locus associated with anomalies of the thoracic arteries and veins^24^.

### Gene-burden results for arrythmias

Within the arrhythmia phenotype group, we identified 28 exome-wide significant gene– phenotype associations, comprising 13 established and 15 novel associations (Supplementary Table 6, Figure 2B). The established associations involved *ACTC1, C10orf71, ENTREP1, KDM5B, LMNA, MYBPC3, MYZAP, PKP2, PLEC, RPL3L, SCN5A, SMADC,* and *TTN*.

A burden of AAVC-classified P or LP variants in *EHMT2* was associated with increased risk of atrial fibrillation (OR = 4.03, P = 3.32 × 10^−10^, λ = 1.03). Qualifying variants were identified in 37 cases and 145 controls. The direction of effect was consistent across the Central and South Asian, European, and other ancestry groups, whereas no qualifying variants were observed in the Admixed American or East Asian groups. *EHMT2* encodes G9a, a histone lysine methyltransferase involved in the epigenetic regulation of gene expression. A recent study identified seven de novo heterozygous *EHMT2* variants, five missense variants and two in-frame deletions, that cause an autosomal dominant *EHMT2*-related Kleefstra syndrome. These variants clustered at the G9a enzyme–substrate interface, and cardiovascular abnormalities were reported in all seven patients^25^. In our analysis, 30 distinct qualifying *EHMT2* variants were observed among the 37 cases, comprising 18 high-impact variants and 12 missense variants. Notably, all of the missense variants were located within the SET domain, which contains the catalytic region of G9a. One of these variants, p.Ala1077Thr, has also been reported in an individual with *EHMT2*-related Kleefstra syndrome. The observation of the same variant in individuals with different phenotypic presentations may reflect incomplete penetrance, variable expressivity, or the influence of additional genetic or environmental modifiers (Supplementary Table 7).

In our analysis, two genes encoding purinergic receptors were associated with arrhythmia phenotypes: *P2RX1*, encoding the P2X1 receptor, and *P2RY4*, encoding the P2Y4 receptor. High-impact variants in *P2RX1* were associated with the combined all-arrhythmia phenotype (OR = 7.13, P = 1.82 × 10^−6^, λ = 1.01), whereas *P2RY4* P+LP+VUS-H variants combined with predicted damaging missense variants were associated with complete atrioventricular block (OR = 2.05, P = 2.00 × 10^−6^, λ = 0.97). P2X1 is an ATP-gated ion channel expressed in the human heart, including atrial tissue, as well as in vascular smooth muscle and platelets. P2Y4 is a G-protein-coupled nucleotide receptor. In the heart, P2Y4 is expressed in cardiac endothelial cells, and loss of the receptor impairs endothelial-cell proliferation and migration, resulting in decreased microvascular and cardiomyocyte proliferation during postnatal cardiac development^26^.

The remaining 12 novel gene–phenotype associations were *ENKD1, RBM12, RC3H2* and *UBR3* with the combined all-arrhythmias; *OR51B5* with right bundle branch block; *MYO1C* with tachycardia; *MDM1* with ventricular fibrillation; *ECI2* with premature beat*s; AGO2, MYH15, PMP2,* and *PPARGC1A* with atrial fibrillation.

### Gene-burden results for cardiomyopathies and heart failures

Twelve genes reached exome-wide significance in the gene-burden analyses for cardiomyopathies and heart failures. Eight represented replications of established gene–phenotype associations*: ALPK3, DSP, GLA, LMNA, MYBPC3, MYH7, TTN,* and *TTR* (Figure 2C and Supplementary Table 8).

*TXNIP* encodes thioredoxin-interacting protein, a regulator of cellular redox signalling, oxidative-stress responses, and metabolism. AAVC-classified P+LP *TXNIP* variants was associated with increased odds of left heart failure in male UK Biobank participants (OR = 6.76, P = 2.81 × 10^−7^, λ = 0.99). All qualifying variants were heterozygous and were identified in 13 cases and 65 controls. In the Mouse Genome Informatics database, homozygous deletion of *Txnip* was associated with abnormal left ventricular morphology, including an increase in left ventricular dimensions, as well as reduced left ventricular fractional shortening. In addition, a recent study describing nine individuals with biallelic pathogenic *TXNIP* variants reported cardiomyopathy in three individuals^27^.

*UBR3* encodes an E3 ubiquitin-protein ligase that is ubiquitously expressed across human tissues. In our analysis, a burden of high-impact variants in *UBR3* was associated with heart failure in the combined-sex analysis (OR = 4.21, P = 3.43 × 10^−7^, λ = 1.00) and, interestingly, with the combined arrhythmia phenotype in males (OR = 5.34, P = 2.07 × 10^−6^, λ = 1.01). UBR3 knockdown increased protein levels of the cardiac voltage-gated sodium channel Nav1.5, encoded by *SCN5A*^28^, and also increased the abundance of the L-type calcium channel Cav1.2, encoded by *CACNA1C*, resulting in increased L-type calcium current and enhanced sarcoplasmic-reticulum Ca²⁺ release in ventricular cardiomyocytes^29^. Notably, gain-of-function variants in *SCN5A* and *CACNA1C* are established causes of cardiac arrhythmia syndromes. Certain variants in these genes have also been associated with dilated and hypertrophic cardiomyopathy, respectively. Additionally, in a previous biobank-scale meta-analysis, rare loss-of-function variants in *UBR3* were associated with hypertension and type 2 diabetes and obesity^9^.

The remaining two novel gene–phenotype associations were *OR4D10* with hypertrophic cardiomyopathy and *ZNFC00* and heart failure.

### Gene-burden results for heart valve disorders

We replicated two of the previously established gene–phenotype associations for heart valve disorders: *FBN1* and *SMADC*. We also identified six novel candidate gene– phenotype associations (Figure 2D, Supplementary Table 9).

The novel associations included *B3GAT2* with aortic insufficiency; *IL1C* and *TM7SF3* with mitral valve insufficiency; and *CD27C* and *RAMAC* with the combined heart valve disorder phenotype. Interestingly, a burden of high-impact variants in *KLLN* was associated with reduced odds of the combined heart valve disorder phenotype across all tested genetic ancestry groups (OR = 0.59, P = 2.09 × 10^−6^, λ = 0.99).

*CD27C* encodes B7-H3, a transmembrane immunoregulatory protein of the B7 family. In our study, a burden of P/LP/VUS-H variants in *CD27C* was associated with increased odds of the combined heart valve disorder phenotype (OR = 1.81, P = 2.45 × 10^−7^, λ = 0.96), with qualifying variants observed in 104 cases and 1,533 controls. *CD27C* has previously been shown to be differentially expressed during postnatal maturation of the aortic valve^30^. More recently, a large GWAS of mitral valve prolapse identified a common variant in *CD27C*^31^. In addition, previous experimental studies have shown that *CD27C* regulates endothelial cell proliferation, migration, and angiogenic differentiation^32^.

### Gene-burden results for embolism and thrombosis

For embolism and thrombosis, we replicated five of the previously established gene– phenotype associations: *F11, PROC, PROS1, SERPINC1,* and *STAB2*. We also identified four novel gene–phenotype associations (Figure 2E, Supplementary Table 10).

A burden of AAVC-classified pathogenic, likely pathogenic, and high-confidence variants of uncertain significance, together with predicted damaging missense variants, in *PIK3C3* was associated with increased odds of phlebitis and thrombophlebitis in female UK Biobank participants (OR = 2.39, P = 1.65 × 10^−6^, λ = 0.92). *PIK3C3*, also known as *VPS34*, encodes the catalytic subunit of the class III phosphatidylinositol 3-kinase complex and regulates autophagy and intracellular trafficking. In platelet- and megakaryocyte-specific mouse models, loss of *Pik3c3* impaired platelet granule secretion, activation, and thrombus growth under arterial flow conditions and reduced thrombus formation after arterial injury^33,34^.

The remaining three novel gene–phenotype associations were *LAMB4* and *SOX15* with pulmonary embolism; and *MAFA* with arterial embolism and thrombosis.

### Gene-burden results for cerebrovascular diseases

In our study, nine genes reached the exome-wide significance threshold, of which three represented established associations: *COL4A2*, *HTRA1*, and *NOTCH3* (Supplementary Table 11, Figure 3A).

**Figure 3:**
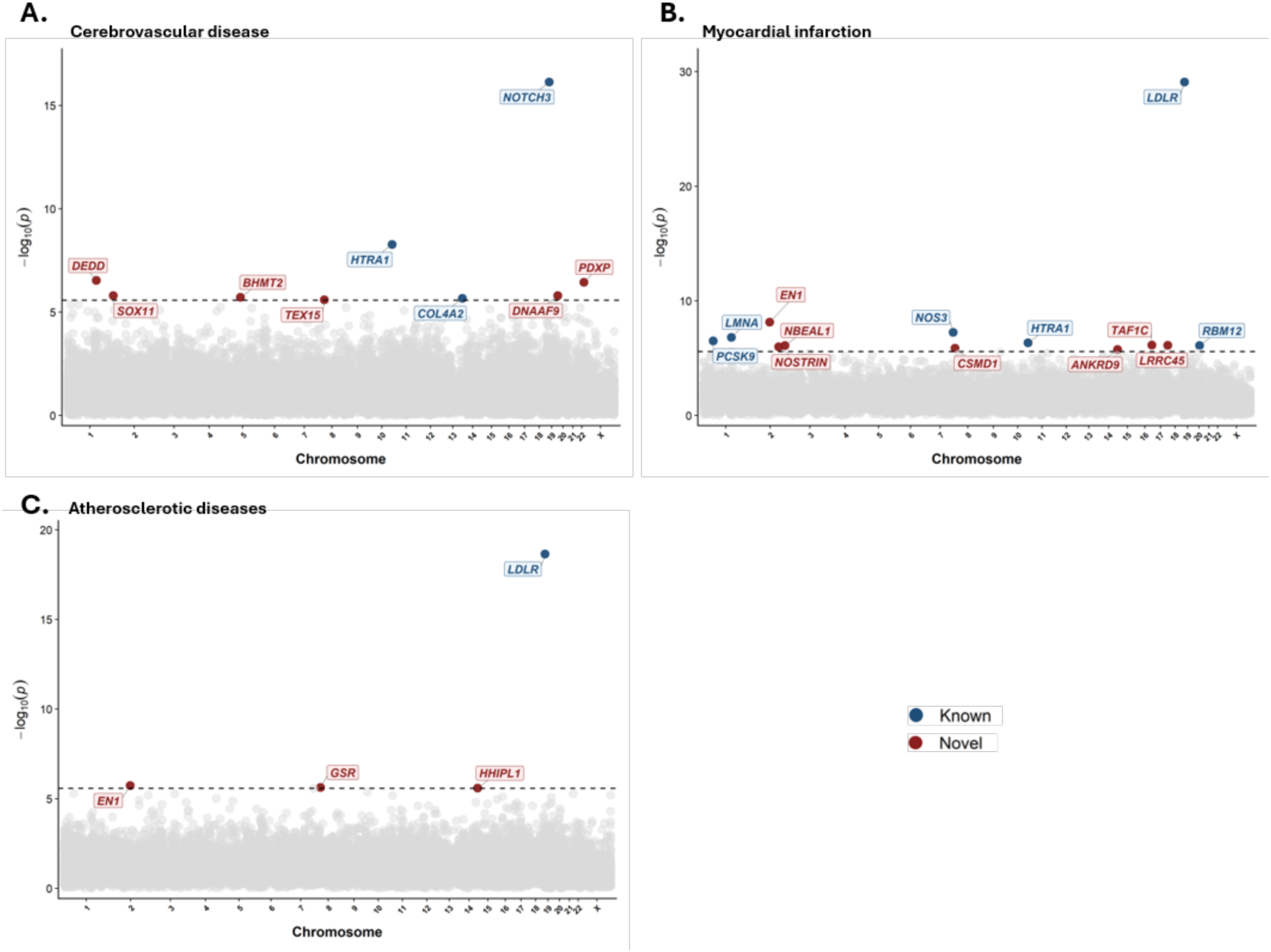
Manhattan plots for gene-based association results for atherosclerotic cardiovascular phenotype groups. Each point corresponds to a gene mapped to its genomic position along chromosomes 1–22 and X, with association significance displayed as −log₁₀(P) on the vertical axis. For each gene–phenotype group combination, the most significant P-value was selected from the eligible association tests, requiring at least 10 qualifying-variant carriers among cases. The dashed horizontal line denotes the Bonferroni-corrected significance threshold (P = 0.05/18,834). Previously established gene–phenotype associations are shown in dark blue, while novel associations are indicated in dark red.

A burden of damaging missense variants in *DEDD* was associated with the combined stroke phenotype in females (OR = 7.66, P = 2.89 × 10^−7^, λ = 0.99), with qualifying variants observed in 12 cases and 35 controls. *DEDD* encodes death effector domain-containing protein, a ubiquitously expressed protein involved in apoptotic signalling. *DEDD* is also highly constrained against genetic variation, with a missense Z score of 3.32 and a LOEUF of 0.27.

A burden of damaging missense variants in *BHMT2* was associated with the combined stroke phenotype (OR = 1.47, P = 1.94 × 10⁻⁶, λ = 1.01), with qualifying variants observed in 191 cases and 2,336 controls. The direction of effect was consistent across African, Central and South Asian, European, and other genetic ancestry groups. *BHMT2* encodes betaine-homocysteine S-methyltransferase 2, a zinc-dependent enzyme involved in homocysteine remethylation. Notably, common *BHMT2* variants have previously been associated with early-onset ischemic stroke^35^. Consistent with the role of BHMT2 in homocysteine metabolism, elevated circulating homocysteine has been associated with an increased risk of both stroke and ischemic stroke in a meta-analysis of prospective cohort studies^36^.

The remaining four novel associations involved *DNAAFS*, *PDXP*, and *SOX11* with ischemic stroke, and *TEX15* with cerebral infarction.

### Gene-burden results for myocardial infarction

In our analysis, we replicated six previously reported associations and identified seven novel myocardial infarction-related genes. The established associations involved *HTRA1, LDLR, LMNA, NOS3, PCSKS,* and *RBM12*, which have previously been implicated in myocardial infarction or coronary artery disease (Supplementary Table 12, Figure 3B).

A burden of AAVC-classified P/LP variants in *NBEAL1* was associated with coronary atherosclerosis in males (OR = 1.72, P = 8.00 × 10^−7^, λ = 0.99). Qualifying variants were observed in 136 cases, including one homozygous carrier, and 575 controls. *NBEAL1* encodes neurobeachin-like protein 1 and is highly expressed in arterial tissues. Common variants at the *NBEAL1* locus have previously been associated with increased risk of coronary artery disease and reduced NBEAL1 expression in arteries^37^. End-truncating high-impact variants in *NOSTRIN* was associated with atherosclerotic heart disease in females (OR = 5.35, P = 1.04 × 10^−6^, λ = 0.97), with qualifying variants observed in 16 cases and 99 controls. *NOSTRIN* encodes nitric oxide synthase trafficking protein and regulates the subcellular localization and activity of endothelial nitric oxide synthase. Deletion of *Nostrin* in endothelial cells reduced nitric oxide and cGMP levels, impaired acetylcholine-induced vascular relaxation, and increased systemic blood pressure in mice^38^.

The remaining five novel associations involved *ANKRDS* with ischemic heart disease, *CSMD1* with ischemic cardiomyopathy, *EN1* with the combined acute myocardial infarction phenotype, *LRRC45* with angina pectoris, and *TAF1C* with coronary atherosclerosis/atherosclerotic heart disease.

### Gene-burden results for atherosclerotic phenotypes

In addition to the separate ischemic stroke and coronary heart disease phenotype groups, we analysed atherosclerosis as a distinct phenotype. We identified three novel exome-wide significant associations involving *EN1*, *GSR*, and *HHIPL1*, and replicated the established association with *LDLR* (Supplementary Table 13, Figure 3C).

A burden of damaging missense variants in *GSR* was associated with atherosclerosis of the aorta (OR = 3.12, P = 2.37 × 10^−6^, λ = 0.95). *GSR* encodes glutathione-disulfide reductase, which maintains cellular glutathione in its reduced form and takes place in oxidative stress response. Glutathione reductase activity has previously been shown to be markedly reduced in human carotid atherosclerotic plaques compared with nonatherosclerotic arteries^39^. In addition, increased expression of glutathione reductase in macrophages reduced atherosclerosis in *Ldlr*-deficient mice without affecting plasma cholesterol or triglyceride levels^40^.

A cumulative burden of AAVC-classified P+LP+VUS-H and damaging missense variants in *HHIPL1* was associated with the combined atherosclerotic disease phenotype (OR = 1.27, P = 2.52 × 10^−6^, λ = 1.02), observed in 598 cases and 2613 controls. *HHIPL1* encodes hedgehog-interacting protein-like 1, a secreted protein that positively regulates Hedgehog signalling. *HHIPL1* lies within the chromosome 14q32 region corresponding to the coronary heart disease susceptibility 4 locus (OMIM: 608318). Experimental studies have shown that HHIPL1 promotes vascular smooth muscle cell proliferation and migration, while *Hhipl1* deficiency reduced atherosclerotic plaque burden by more than 50% in both *Apoe*- and *Ldlr*-deficient mice^41^.

### Sensitivity analysis with cardiometabolic traits

We conducted sensitivity analyses across all cardiovascular phenotypes with cardiometabolic traits used as covariates. Of the 40 replicated gene–phenotype associations, 34 (85%) remained exome-wide significant after adjustment, while 26 of the 46 (57%) novel associations retained exome-wide significance (Figure 4A, 4B, 4C; Supplementary Table 14 and 15).

**Figure 4:**
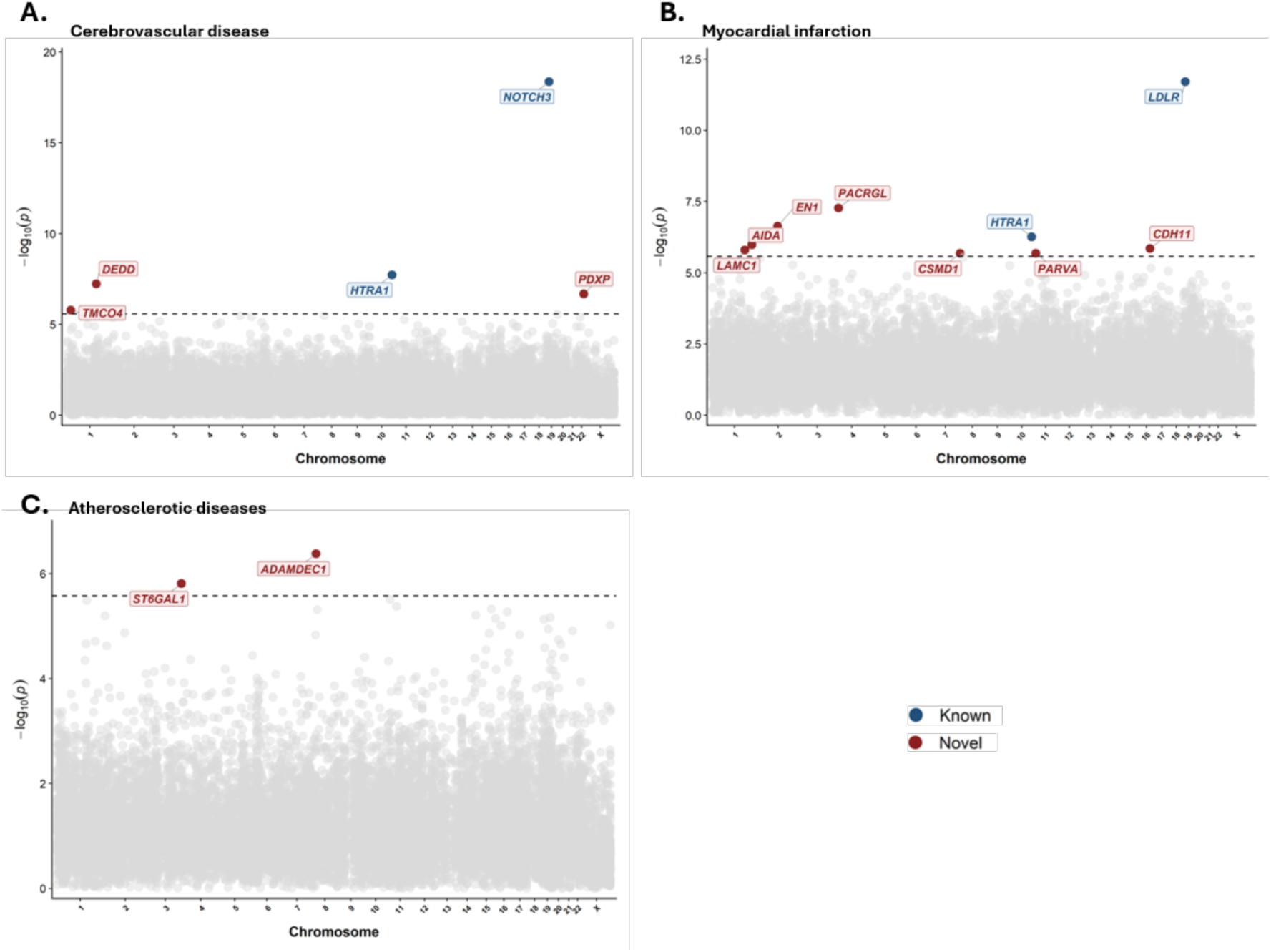
Manhattan plots for gene-based association results for atherosclerotic cardiovascular phenotype groups after the sensitivity analysis. Each point denotes a gene plotted according to its chromosomal position (1–22 and X), with association significance expressed as −log₁₀(P) on the vertical axis. The lowest P-value was retained for each gene– phenotype group combination across eligible association tests, restricted to those with at least 10 qualifying-variant carriers among cases. The horizontal dashed line marks the Bonferroni-corrected significance threshold (P = 0.05/18,834). Established gene–phenotype associations are indicated in dark blue, whereas novel associations are shown in dark red.

The effect of cardiometabolic adjustment differed substantially between atherosclerosis-related and non-atherosclerotic phenotypes. Among replicated associations involving atherosclerosis-related phenotypes, including cerebrovascular disease, myocardial infarction, and atherosclerosis, 40% remained exome-wide significant after adjustment. In contrast, all replicated associations involving non-atherosclerotic phenotypes retained exome-wide significance. A similar pattern was observed among the 46 novel associations: only 25% (4/16) of associations involving atherosclerosis-related phenotypes remained exome-wide significant after cardiometabolic adjustment, compared with 73% (22/30) of associations involving non-atherosclerotic phenotypes (Supplementary Table 16).

In the sensitivity analysis, an additional 18 gene–phenotype associations reached exome-wide significance (Supplementary Table 15). These associations included *TPTE* with aortic aneurysm and dissection; *ERCC5, MST1R, SERINC5, TXLNG,* and *TYMS* with arrhythmia phenotypes; *ADAMDEC1* and *STCGAL1* with atherosclerosis; *TRIM10* and *ZNFC15* with cardiomyopathy and heart failure; *TMCO4* with cerebrovascular disease; *AIDA, CDH11, LAMC1, PACRGL,* and *PARVA* with myocardial infarction; and *TAS1R3* and *ZUP1* with heart valve disease.

In the sensitivity analysis, a burden of P+LP+VUS-H variants in *CDH11* reached exome-wide significance with ischemic heart disease after adjustment for cardiometabolic traits (OR = 4.40, P = 1.41 × 10^−6^, λ = 1.03), with qualifying variants observed in 24 cases and 110 controls in the sensitivity analysis. *CDH11* encodes cadherin-11, a calcium-dependent cell–cell adhesion protein involved in cellular adhesion, migration, inflammation, and tissue remodeling. Biallelic loss-of-function variants in *CDH11* cause autosomal recessive Elsahy–Waters syndrome (OMIM: 211380), whereas heterozygous missense variants in *CDH11* cause Teebi hypertelorism syndrome 2 (OMIM: 619736). In previous studies, in an *Apoe*-deficient mouse model of atherosclerosis, *Cdh11* expression was increased approximately threefold in atherosclerotic plaques and correlated with markers of macrophage infiltration and inflammation. Additionally, loss of *Cdh11* resulted in increased plaque burden and vascular inflammation and altered macrophage migration and T-cell responses^42^.

A combined burden of P+LP+VUS-H variants in *PARVA* was associated with atherosclerotic heart disease after adjustment for cardiometabolic traits (OR = 2.51, P = 2.09 × 10^−6^, λ = 1.02), with qualifying variants observed in 50 cases and 277 controls. *PARVA* encodes α-parvin, a focal-adhesion adaptor protein that forms part of the integrin-linked kinase–PINCH–parvin complex and links integrin-mediated adhesion to the actin cytoskeleton. Previous experimental studies in mice demonstrated that loss of α-parvin impaired the recruitment of vascular smooth-muscle cells and pericytes to developing vessels, disrupted vessel remodelling, and increased vascular smooth-muscle-cell contractility^43^. Endothelial-specific loss of α-parvin similarly impaired angiogenesis and vascular stability by disrupting endothelial cell–cell junctions, resulting in increased vessel regression and hemorrhage^44^.

A burden of singleton damaging missense variants in *ADAMDEC1* was associated with the combined atherosclerotic phenotype (OR = 11.07, P = 4.18 × 10^−7^, λ = 0.96) in females, reaching exome-wide significance only in the sensitivity analysis after adjustment for cardiometabolic traits. *ADAMDEC1* encodes ADAM-like decysin-1, a secreted metalloprotease expressed predominantly by macrophages and dendritic cells and involved in immune and inflammatory processes. In previous studies ADAMDEC1 expression was increased in human atherosclerotic samples and experimental atherosclerosis models, while *ADAMDEC1* knockdown reduced atherosclerotic lesion formation and attenuated vascular-cell responses to oxidized LDL^45^. More recently, ADAMDEC1 knockdown was shown to reduce aortic lesion burden and suppress vascular smooth-muscle-cell proliferation and migration^46^. Given that reduced ADAMDEC1 activity appears protective in experimental models, whereas the positive association in our study was driven by rare damaging missense variants, we hypothesize that some of these variants may exert altered-or gain-of-function effects on ADAMDEC1 activity.

### Penetrance estimates and genetic prevalence for cardiovascular disease– associated genes

We leveraged the gene-burden summary statistics to estimate the penetrance of cardiovascular phenotypes among carriers of qualifying variants for each gene– phenotype association. Estimated penetrance varied substantially across associations, ranging from 53% for *ENKD1* with the combined arrhythmia phenotype to 0.5% for *TTN* with dilated cardiomyopathy (Supplementary Table 17). We observed an inverse relationship between penetrance and the number of qualifying variant carriers in the UK Biobank, with higher-penetrance gene–phenotype associations represented by fewer carriers (R² = 0.54, P = 5.23 × 10^−16^), which is consistent with population-genetic expectations that highly penetrant disease-causing variation is constrained to lower population frequencies^47^ (Figure 5), as well as with observations across monogenic disease genes showing substantial differences in penetrance and carrier frequency for the same phenotype^48^.

**Figure 5:**
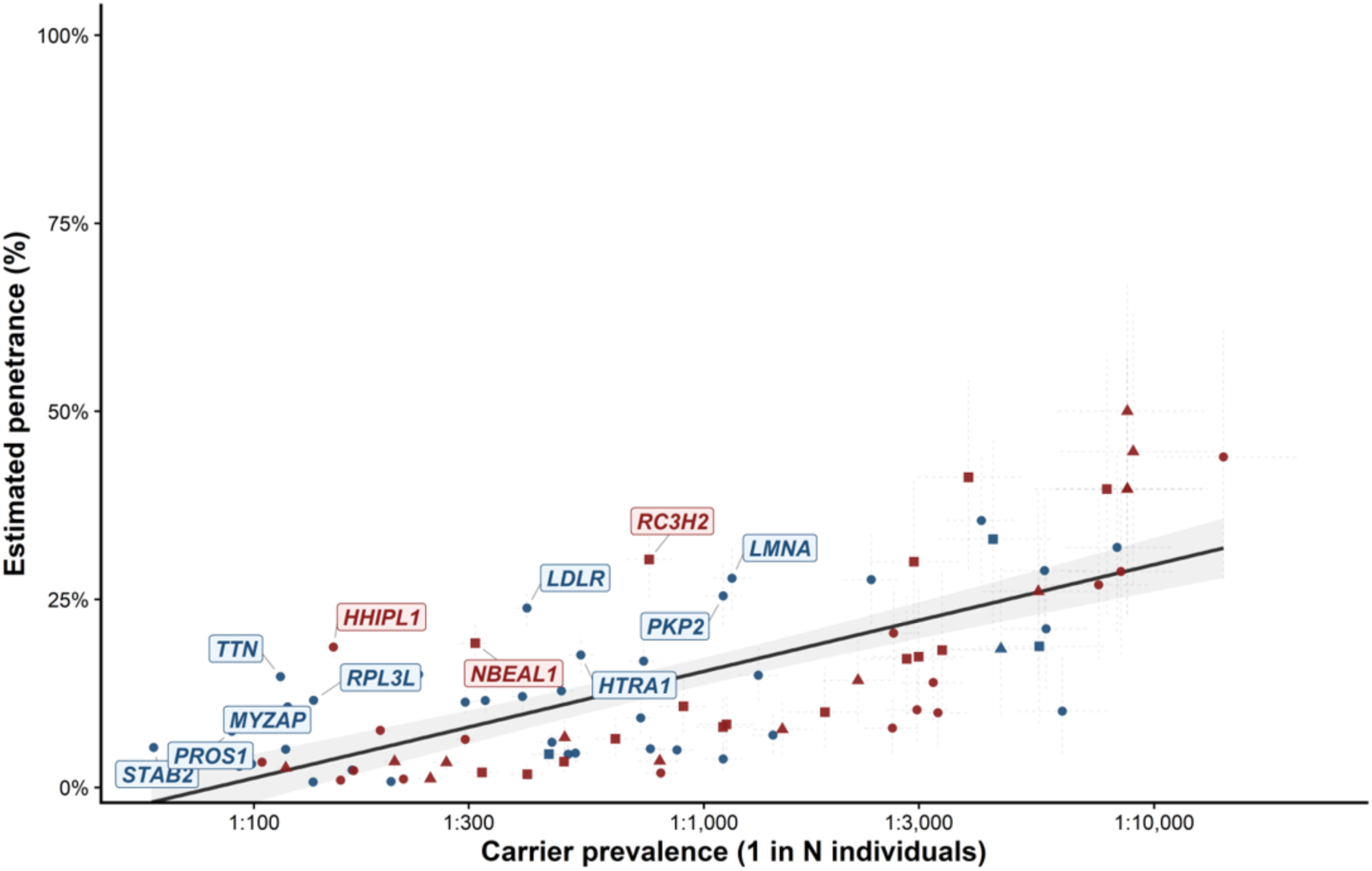
Relationship between carrier frequency and estimated penetrance of cardiovascular disease-associated genes. Each point represents a gene–phenotype association, with the horizontal axis indicating the prevalence of qualifying-variant carriers, expressed as one carrier per N individuals on a logarithmic scale, and the vertical axis showing estimated penetrance (%). Horizontal dashed error bars represent exact 95% confidence intervals for carrier prevalence, whereas vertical dashed error bars represent 95% Bayesian credible intervals for penetrance. The solid black line indicates the fitted unweighted linear regression of estimated penetrance on log₁₀-transformed carrier prevalence. Previously established associations are shown in dark blue, novel associations in dark red. Point shapes distinguish combined-sex, female-specific, and male-specific analyses. Genes confidently exceeding the fitted regression model for both prevalence and penetrance estimates are labelled.

Next, we compared penetrance estimates and association statistics across different variant masks for the same gene–phenotype associations. For example, both the P variants mask and the broader P+LP+VUS-H variant mask in *SCN5A* were associated with the combined arrhythmia phenotype. Qualifying variants were observed in 286 and 1744 individuals, respectively. Estimated penetrance was higher for P variants (24.8%) than for the broader P+LP+VUS-H mask (19.2%). In contrast, the statistical evidence for association was stronger for the more inclusive P+LP+VUS-H mask (P = 1.27 × 10^−9^) than for P variants alone (P = 3.12 × 10^−7^). Across all gene–phenotype associations for which both a broader and a more stringent variant mask were significant; we identified 77 paired comparisons. The more stringent mask yielded a more significant association P value in 41 of the 77 comparisons. In contrast, the more stringent mask showed a higher estimated penetrance in 73 of 77 comparisons, indicating that restricting the analysis to more confidently deleterious variants generally increased penetrance estimates, even when it did not consistently strengthen the statistical evidence for association (Supplementary Table 18).

For genetic prevalence estimation, we used AAVC to classify variants reported in gnomAD in both established cardiovascular disease genes and the novel genes identified in this study. We then estimated the prevalence of pathogenic and likely pathogenic genotypes across eight gnomAD genetic ancestry groups: African/African American, Admixed American, Ashkenazi Jewish, East Asian, Finnish, Middle Eastern, non-Finnish European, and South Asian^49^.

The estimated prevalence of qualifying genotypes varied substantially across cardiovascular phenotype groups. For aortic aneurysm and dissection, approximately 1 in 62 individuals carried a qualifying genotype, with the largest gene-specific contributions from the novel candidate *TPTE* (1 in 194), followed by *SMADC* (1 in 791) and *THSD4* (1 in 826). Arrhythmia-associated genotypes were considerably more frequent, occurring in approximately 1 in 8.9 individuals; the most prevalent were in *HFE* (1 in 64), *TTN* (1 in 97), and the novel candidate *MYH15* (1 in 167). Similarly, approximately 1 in 10 individuals carried a qualifying genotype associated with cardiomyopathy or heart failure, with *HFE* and *TTN* again contributing most prominently, followed by *CPT2* (1 in 149). For cerebrovascular disease, the estimated genetic prevalence was approximately 1 in 17 individuals, with the most prevalent qualifying genotypes occurring in *ABCCC* (1 in 90), the novel candidate *TEX15* (1 in 133), and *RNF213* (1 in 171). Approximately 1 in 26 individuals carried a qualifying genotype associated with myocardial infarction, with the largest contributions from *LDLR* (1 in 150), *APOE* (1 in 164), and the novel candidate *NBEAL1* (1 in 208). For embolism and thrombosis, qualifying genotypes were present in approximately 1 in 13 individuals and were most frequent in *F5* (1 in 31), *F11* (1 in 71), and *STAB2* (1 in 161). For heart valve disorders, approximately 1 in 14 individuals carried a qualifying genotype, with *ABCCC* (1 in 90), *LPA* (1 in 199), and *SCN5A* (1 in 225) showing the highest gene-specific prevalence (Figure 6A:E, Figure 7A:B and Supplementary Table 19 and 20).

**Figure 6:**
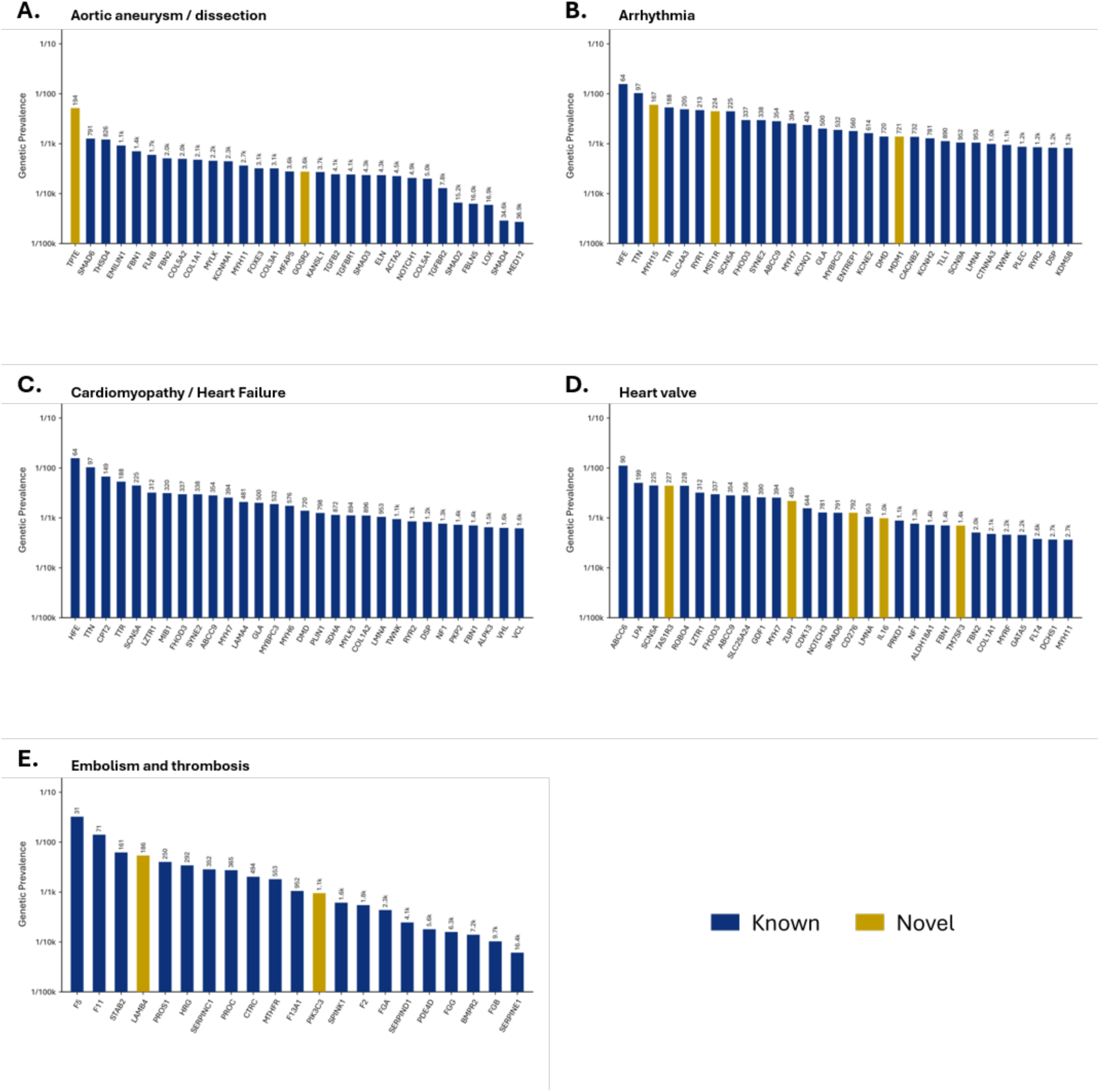
Genetic epidemiology of genes associated with non-atherosclerotic cardiovascular phenotypes. Estimated genetic prevalence of pathogenic and likely pathogenic variant genotypes in genes associated with non-atherosclerotic cardiovascular phenotypes. Qualifying variants were classified using AAVC, and allele-frequency information was obtained from eight genetic ancestry groups represented in gnomAD.

**Figure 7:**
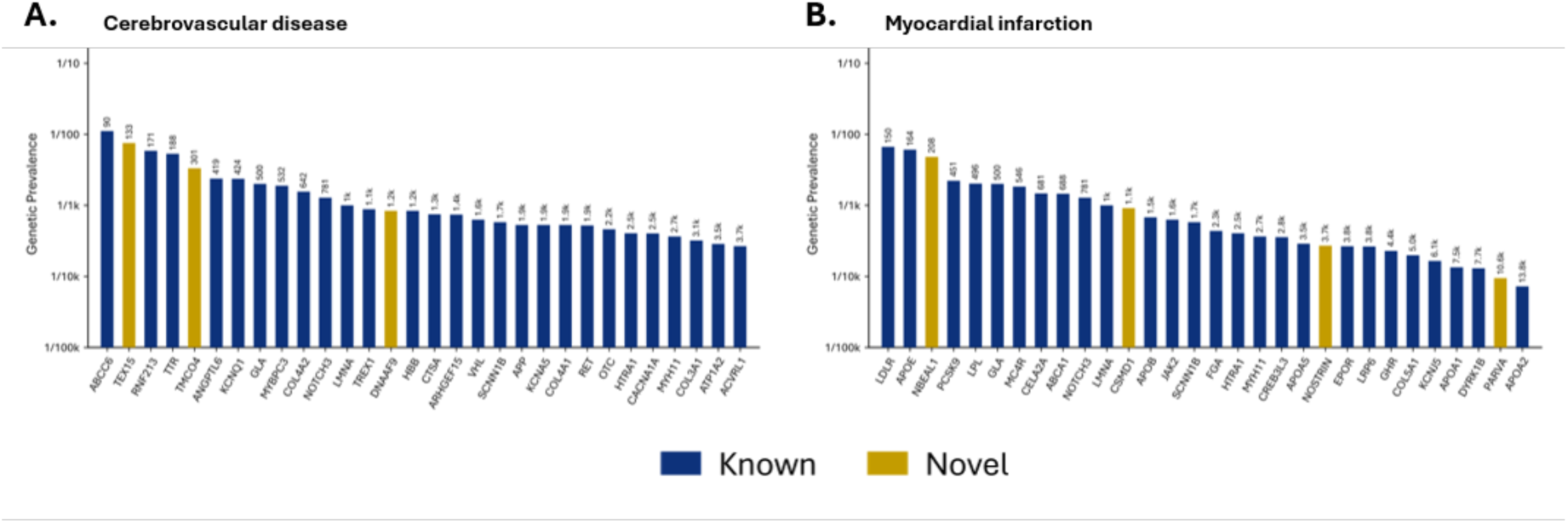
Genetic epidemiology of genes associated with atherosclerotic cardiovascular phenotypes. Genetic prevalence was estimated for genes associated with atherosclerotic cardiovascular phenotypes using variants classified as pathogenic or likely pathogenic by AAVC. Allele-frequency data were obtained from eight genetic ancestry groups included in gnomAD.

Across all cardiovascular phenotype groups, approximately 1 in 3.5 individuals carried at least one qualifying P/LP genotype in an established or novel cardiovascular disease-associated gene (Supplementary Table 20).

## Discussion

We performed large-scale gene-based rare-variant analyses across eight cardiovascular phenotype groups using whole-genome sequencing data from up to 483,311 UK Biobank participants, incorporating sex-stratified analyses, multiple ancestry groups, and variant masks spanning different levels of ACMG classifications and variant effects. We identified 86 exome-wide significant gene–phenotype associations, including 40 established and 46 novel relationships. Additionally, we performed sensitivity analyses to assess the extent to which the observed cardiovascular gene–phenotype associations were influenced by cardiometabolic risk factors.

Purinergic signalling can influence cardiac electrophysiology, vascular function, and endothelial–myocardial interactions, indicating that disruption of extracellular nucleotide signalling may contribute to arrhythmogenesis through multiple, potentially complementary, cellular mechanisms. Two purinergic receptor genes, *P2RX1* and *P2RY4*, were associated with arrhythmia phenotypes, suggesting a potential role for extracellular nucleotide signalling in arrhythmia susceptibility. In previous experimental studies, extracellular ATP induced delayed after-depolarizations, ectopic activity, and ventricular tachycardia, effects that were attenuated by P2-receptor antagonists^50^.

We identified an association between end-truncating *NOSTRIN* variants and atherosclerotic heart disease in females, implicating an additional gene involved in endothelial nitric oxide signalling. *NOSTRIN* encodes nitric oxide synthase trafficking inducer, which directly interacts with endothelial nitric oxide synthase, encoded by *NOS3*, an established coronary artery disease gene^51^, and regulates its subcellular localization and nitric oxide production^52^. In experimental studies, endothelial-specific deletion of *Nostrin* in mice was previously shown to reduce nitric oxide and cGMP levels, impair acetylcholine-induced vascular relaxation, and increase systemic blood pressure^38^. Similarly, rare inactivating variants in *NOS3* or *GUCY1A3*, which encodes a component of the downstream nitric oxide receptor, were associated with an approximately three-fold increased risk of coronary heart disease^53^. The association between *NOSTRIN* and atherosclerotic heart disease extends the human genetic evidence for this pathway by implicating a regulator of eNOS trafficking and activity, further supporting the contribution of endothelial nitric oxide homeostasis to coronary atherosclerotic disease.

Our companion cardiometabolic study connects rare coding variation to metabolic and vascular risk-factor phenotypes^18^, providing a physiological context for the sensitivity analyses. We observed that associations involving atherosclerosis-related phenotypes less often retained exome-wide significance after adjustment for conventional cardiometabolic risk factors than those involving non-atherosclerotic phenotypes. However, several associations, including *CDH11*, *PARVA*, and *ADAMDEC1*, reached exome-wide significance only after cardiometabolic adjustment. Interestingly, all three genes have experimental evidence supporting roles within the vascular wall^42–46^. The known biological functions prioritize investigation of vascular inflammation, endothelial integrity, smooth-muscle-cell behaviour, and vessel remodelling as mechanisms linking these genes to vascular disease.

Our findings broaden the cardiovascular spectrum of genes implicated in recessive disorders. *EN1*, implicated in limb–brain malformations, was associated with myocardial infarction and atherosclerosis in our study; *GOSR2*, linked to progressive myoclonus epilepsy and congenital muscular dystrophy, with aortic aneurysm. Associations of *GSR* with aortic atherosclerosis and *TEX15* with cerebral infarction extend beyond their established diseases of glutathione reductase deficiency and spermatogenic failure, respectively. *TXNIP*, already implicated in metabolic disease and cardiomyopathy in biallelic form, was associated with left heart failure in heterozygous males. These observations highlight allele dosage, molecular consequence and tissue context.

Comparisons across variant masks further showed that statistical significance and observed penetrance are not equivalent. Among 77 paired comparisons in which both masks reached exome-wide significance, the more stringent mask had higher estimated penetrance in 73, but a more significant P value in only 41. Broader masks may therefore improve statistical power by increasing carrier numbers, whereas more confidently pathogenic variants generally identify carrier groups with higher observed penetrance.

Across the reported gene–phenotype combinations, penetrance was inversely related to carrier frequency. This distinction is important when interpreting genetic-prevalence estimates. We estimated that approximately one in 3.5 individuals carry at least one qualifying P/LP genotype across the cardiovascular gene set. However, this should not be interpreted as an equivalent prevalence of cardiovascular disease, as; disease expression depends on penetrance, variant-specific effects, age, genetic background, and environmental exposures.

The newly implicated genes broaden the genomic landscape of inherited susceptibility to cardiovascular disease and offer avenues for mechanistic studies and therapeutic exploration. Our findings may also inform presymptomatic genetic testing and preventive strategies.

## Data Availability

Gene-phenotype association results produced in this study are available at https://zenodo.org/records/23687128. All other data generated or analysed during this study are included in the Supplementary Tables. We used publicly available softwares: AAVC and FuncVEP are available at https://github.com/OzcelikLab/AAVC and https://github.com/OzcelikLab/FuncVEP. REGENIE and remeta are available at https://rgcgithub.github.io/regenie/ and https://rgcgithub.github.io/remeta/.

## Acknowledgements

We gratefully acknowledge İclal Büyükdevrim Özçelik for her invaluable contributions. This research was conducted using the UK Biobank Resource under application number 268863. This work uses data provided by patients and collected by the NHS as part of their care and support. Illustrations in the graphical abstract were adapted from Servier Medical Art, licensed under the Creative Commons Attribution 4.0 International licence.

## Sources of Funding

This study was supported by a grant from the Leducq Foundation. T.Ö. acknowledges financial support received through membership in the Turkish Academy of Sciences. B.K. and K.Ç. acknowledge fellowship support through the 2211-A National Doctoral Scholarship Programme of the Scientific and Technological Research Council of Türkiye. The UK Biobank application fee was covered by the Global Researcher Access Fund.

## Supplemental Material

Supplementary Information

Tables S1-S20

## Disclosures

The authors declare no competing interests.

## Notes

### Competing Interest Statement

The authors have declared no competing interest.

### Clinical Trial

Not applicable.

### Author Declarations

We follow the UKB IRB rules.

